# Altered dynamic functional connectivity in antagonistic state in first-episode, drug-naive patients with major depressive disorder

**DOI:** 10.1101/2024.07.02.24309338

**Authors:** Min Wang, Tao Chen, Zhongyi He, Lawrence Wing-Chi Chan, Qinger Guo, Shuyang Cai, Jingfeng Duan, Danbin Zhang, Xunda Wang, Yu Fang, Hong Yang

**Affiliations:** Key Laboratory for Biomedical Engineering of Ministry of Education, College of Biomedical Engineering and Instrument Science, Zhejiang University, Hangzhou, China; School of Medicine, Sir Run Run Shaw Hospital, Department of Endocrinology, Zhejiang University, Hangzhou, Zhejiang, China; Department of Radiology, The First Affiliated Hospital, College of Medicine, Zhejiang University, Hangzhou, China; Department of Health Technology and Informatics, Hong Kong Polytechnic University, Hong Kong, China; Department of Psychiatry, The First Affiliated Hospital, College of Medicine, Zhejiang University, Hangzhou, China; Department of Psychiatry & Behavioral Sciences, Stanford University School of Medicine, Stanford, CA, USA

**Keywords:** Major depressive disorder, Dynamic functional connectivity, network antagonism, Resting-state functional magnetic resonance imaging

## Abstract

Major depressive disorder (MDD) is characterized by disrupted functional network connectivity (FNC), with unclear underlying dynamics. We investigated both static FNC (sFNC) and dynamic FNC (dFNC) on resting-state fMRI data from drug-naive first-episode MDD patients and healthy controls (HC). MDD patients exhibited lower sFNC within and between sensory and motor networks than HC. Four dFNC states were identified, including a globally-weakly-connected state, a cognitive-control-dominated state, a globally-positively-connected state, and an antagonistic state. The antagonistic state was marked by strong positive connections within the sensorimotor domain and their anti-correlations with the executive-motor control domain. Notably, MDD patients exhibited significantly longer time dwelling in the globally-weakly-connected state, at the cost of significantly shorter time dwelling in the antagonistic state. Further, only the mean dwell time of this antagonistic state was significantly anticorrelated to disease severity measures. Our study highlights the altered dynamics of the antagonistic state as a fundamental aspect of disrupted FNC in early MDD.

## 1. Introduction

Major depressive disorder (MDD) is a common psychiatric disorder characterized by depressed mood, lack of interest with anhedonia, and reduced energy [1,2], which has been reported to affect approximately 4.4% of the population [3,4]. The underlying mechanisms of MDD have been intensively studied in the past [5], and the disrupted brain network is identified as one of the major characteristics as well as the indicator of their mood and cognitive impairment [6,7]. However, the neurophysiological basis of MDD is still not entirely clear.

Functional MRI (fMRI) has been widely used to reveal the alterations of brain networks. Previous static functional network connectivity (sFNC) studies have reported that patients with MDD exhibit unbalanced communication within higher-order networks such as the default mode network (DMN), executive control network (ECN), salience network (SN) and attention network (ATN), collectively referred as the cognitive-control domain in brain [8–11]. Meanwhile, the lower-order networks such as the somatosensory network (SMN), visual network, and cerebellum, which constitute the sensorimotor domain in brain, were also found impaired [12–15]. Abnormalities in cognitive-control domain are often found to affect cognitive and affective processes [10,16], while dysregulation in sensorimotor domain is thought to be associated with psychomotor retardation in MDD patients [17,18]. Dysfunction in cognitive-control and sensorimotor networks were extensively a concern in MDD, while the crosstalk among multi-scale networks, especially the variation in their antagonistic relationships, was largely unknown.

Much neuroimaging evidence has pointed out a decrease of positive correlations between major networks [9,19,20]. Few studies have focused on the negatively correlated networks in MDD [21–23]. Negative correlations between networks are thought to play an important role in coordinating and competing among information-processing activities [24]. Previous studies on negative correlation changes between networks have not reached a consistent conclusion. Decreased strength of negative correlations was reported within the amygdala region [21], between the hippocampus and the salience and prefrontal-parietal networks in MDD patients [25]. Conversely, enhanced antagonism was found between the left amygdala and the left orbitofrontal cortex [23], and between task-positive and task-negative networks in MDD patients [24]. The lack of consistent evidence for alterations in anti-correlations among predefined brain regions or networks may raises doubts about the antagonistic effect between networks in MDD.

However, sFNC is based on presumed stability throughout the scan period and may not reflect the true time-varying properties of FNC. Using the sliding window approach, dynamic functional network connectivity (dFNC) can identify the time-varying characteristics of functional networks [26]. It not only helps uncover details being averaged out in sFNC but also provides more abundant information about FNC dynamics, capturing spontaneously recurring patterns of FNC (i.e., FNC states), thus facilitating the understanding of true communication pathways inside the brain. Recent studies have reported aberrant dFNC in whole-brain or predefined networks perspective in MDD. For example, patients with MDD have higher temporal variability in the medial prefrontal cortex and cingulate gyrus regions [27,28], as well as higher global synchronization and reduced global metastability, which are associated with ruminative thinking [29]. Studies exploring dFNC states also suggest that MDD patients are characterized by a predominance of weakly connected states, rather than strongly connected states, revealing the state dependence of MDD [7,30]. The dFNC is an in-depth complement to sFNC, if combining sFNC and dFNC, we could better understand mental disorders than sFNC alone.

Hence, the purpose of this study is to explore abnormalities in time-varying brain activity and network dynamics in MDD, and to investigate how network dynamics were shifted in early MDD. Considering the medication effects on brain networks and neuronal activity reported in many previous studies [31,32], the investigation of sFNC/dFNC was conducted between first-episode, drug-naive MDD patients and aged-matched healthy controls in this study. We hypothesize that in first-episode, drug-naive adult patients with major depressive disorder (1) there would be disrupted sFNC pattern in different functional networks; (2) altered dFNC properties should reveal more network differences and details than sFNC alone. (3) the dFNC properties would be linked to the symptoms of MDD.

Here we find the scarcity of antagonistic state between executive-motor control networks and sensorimotor networks in MDD. These temporal properties of the antagonistic state may be potential neural markers for monitoring the occurrence of first-episode MDD and predicting the disease severity. In contrast, MDD subjects tend to stay in the globally-weakly-connected state which may represent a characteristic phenomenon of self-focused rumination and impaired pathway of processing and delivering the outside world information. These disrupted dFNC patterns provide new clues to understanding the neuropathology of state-dependence in MDD.

## 2. Results

### 2.1. Demographics and clinical parameters

The demographic and clinical characteristics are summarized in Table 1. No significant differences were found between the MDD and HC groups in terms of gender (t = 0.230, *p* = 0.818) and age (t = −1.491, *p* =0.139). The total years of education existed the group difference in MDD and HC groups (t = ×3.744, *p*<0.001). The mean illness duration was 32.13 ±45.25 months. The mean score of Hamilton Depression Rating Scale (HAMD) was 22.48 ±4.74, while the mean score of Hamilton Anxiety Scale (HAMA) was 15.38 ±5.21. Seventeen of these patients have the HAMD scores of 25-52, considered as severe depression; 27 of them scored of 18-24, considered as moderate depression while 4 of them scored 13-17, considered as mild depression.

**Table 1.** Demographic and clinical data of depressive patients and healthy controls.

| Variables | All MDD patients | Mild MDD | Moderate MDD | Severe MDD | HC | $\chi^2 / t$ | *p*-value |
| --- | --- | --- | --- | --- | --- | --- | --- |
| Gender(male/female) | 12/36 | 1/3 | 6/21 | 5/12 | 13/35 | 0.230 | 0.818 |
| Age (years) | 25.54±5.96 | 22.50±3.11 | 25.11±4.41 | 26.94±8.14 | 27.65±7.76 | -1.491 | 0.139 |
| Education (years) | 14.56±2.81 | 15.25±2.87 | 15.30±2.27 | 13.24±3.21 | 16.67±2.70 | -3.744 | <0.001** |
| Illness duration (months) | 32.13±45.25 | 15.25±18.32 | 30.59±33.74 | 38.53±63.12 | - | - | - |
| HAMD (score) | 22.48±4.74 | 14.25±1.26 | 20.44±2.22 | 27.65±2.32 | - | - | - |
| HAMA (score) | 15.38±5.21 | 9±4.08 | 13.78±3.54 | 19.41±4.94 | - | - | - |

All quantitative data are expressed as mean ± standard deviation; Abbreviation: MDD = major depressive disorder; HC = healthy controls; HAMD = Hamilton Depression Rating Scale; HAMA = Hamilton Anxiety Scale. *p* values displaying significant differences between HC and all MDD patients are marked with an asterisk, \*\*\**p* < 0.001.

### 2.2. Characteristics of the sFNC

Through ICA analysis, 16 ICs were identified within the nine major brain networks, including auditory network (AUN, IC 30), visual network (VN, IC 15,18,20), somatosensory network (SMN, IC 3,10,29), visuomotor integration network (VMN, IC 45), attention network (ATN, IC 31,63), default-mode network (DMN, IC 54,69), executive control network (ECN, IC 37,44), salience network (SN, IC 40), cerebellum (CB, IC 57), which were all similarly obtained in previous studies [33–35] and the detailed spatial maps were represented in Figure 1.

**Figure 1:**
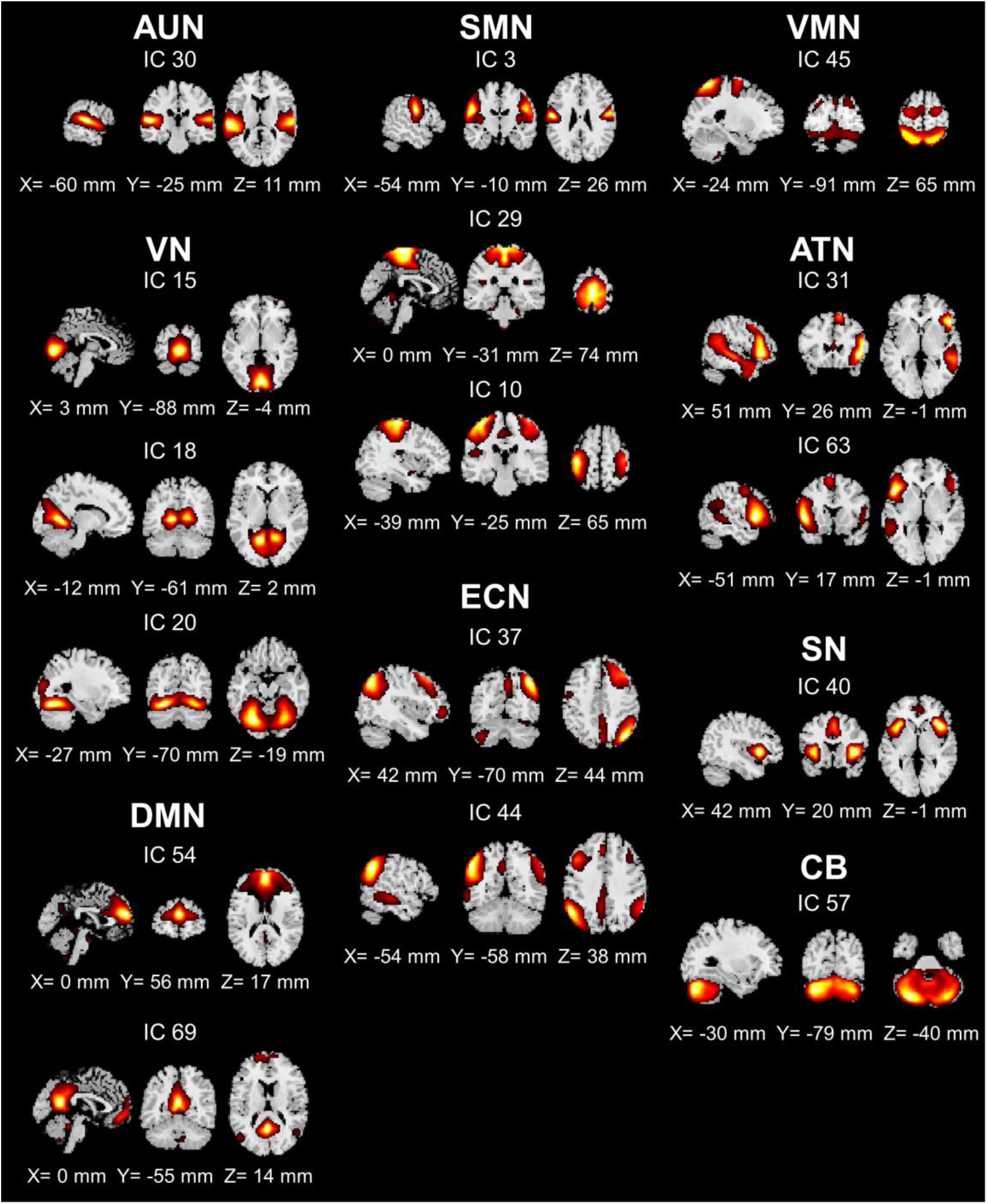
Spatial map of 9 functional networks. Spatial maps of the 16 selected independent components have been identified and grouped into nine functional networks: Auditory network (AUN), visual network (VN), somatosensory network (SMN), default-mode network (DMN), executive control network (ECN), visuomotor integration network (VMN), attention network (ATN, including ventral and dorsal ATN), salience network (SN), cerebellum (CB). The slice location was labeled as the MNI coordinates.

In Figure 2, the cc-matrix was plotted separately for HC and MDD group (Figure 2a, 2b, source data provided in Supplementary Data 1). In both groups, the positive connectivity was found within and in-between the functional networks of AUN, VN, SMN and VMN which in combine act as the sensorimotor domain of the brain. The positive connectivity was also found within and in-between the functional networks of DMN, ECN and ATN, which in combine act as the cognitive-control domain of the brain. Compared to HCs, patients with MDD showed significantly decreased connectivity between SMN and VN, as well as increased connectivity between CB and VN (p<0.05, FDR-corrected) (Figure 2c, 2d).

**Figure 2:**
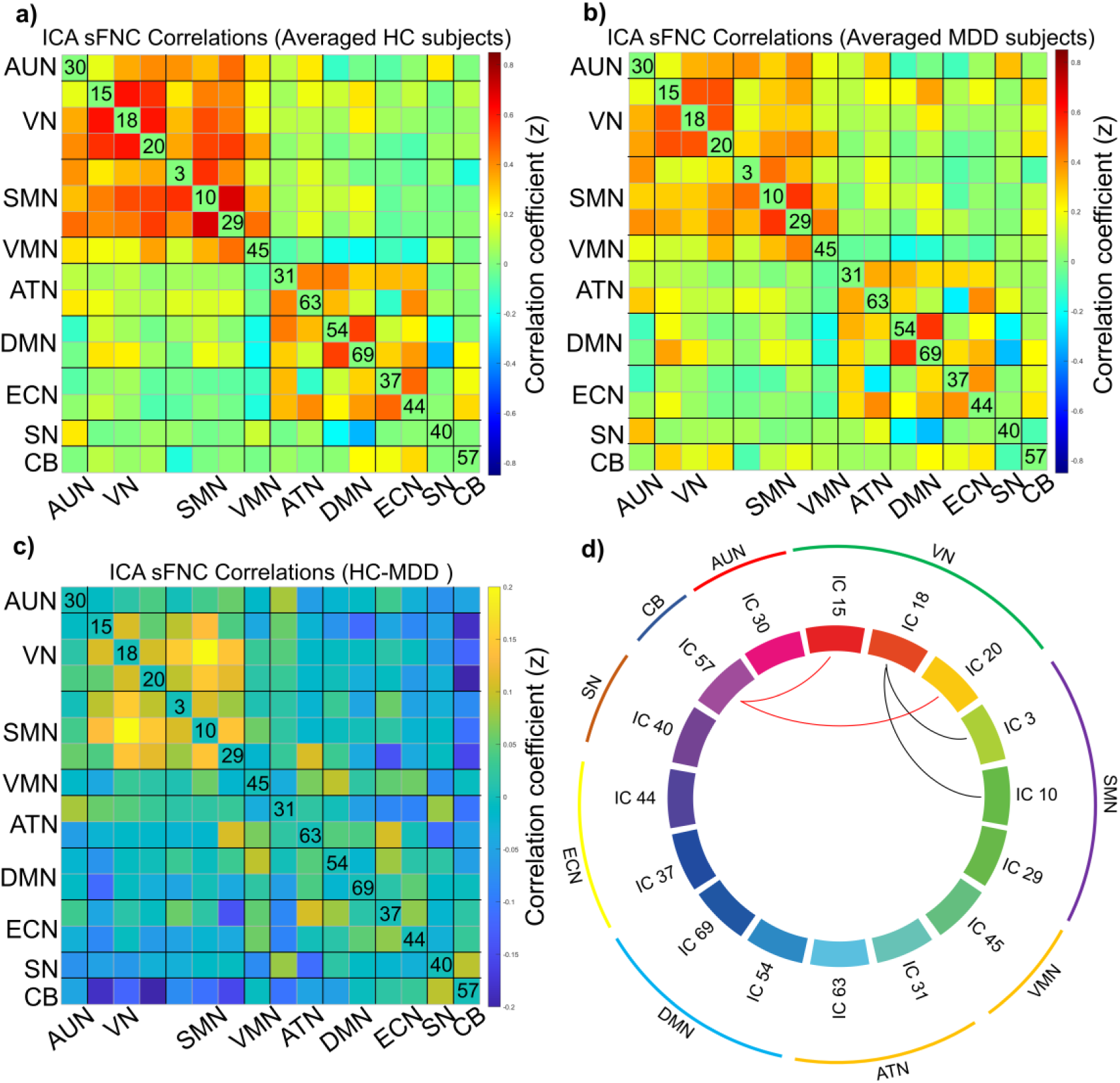
Static matrix of MDD and HC groups and the difference between the two groups. The cc-matrixes were obtained from the ICA time course and have averaged the HC group (a) and MDD group (b). The group difference of cc-matrix was presented in (c) and the significant difference network connectivity was showed in (d, black line: MDD<HC; red line: MDD>HC, p<0.05, FDR-corrected).

### 2.3. Characteristics of the dFNC states

A total of 180 dFNC windows were clustered into four connectivity states that were repeated in all participants. The centroids of the four states were represented in Figure State 1 represented that the whole brain was weakly or sparsely connected. State 1 accounted for the longest time (43%) and represented the sparse FNCs among all networks, 47 HCs and 48 MDDs have entered this state. State 2, which accounts for 24% in all windows, showed positive connectivity within the cognitive-control domain (ATN-DMN-ECN), especially within DMN. State 3 represented strong positive connectivity within the sensorimotor domain (AUN-VN-SMN-VMN) as well as a distinct antagonistic relationship between executive and motor control networks (ECN-CB) and the sensorimotor domain. Thirty-eight HCs and 34 MDDs entered state 3, accounting for 18% of all windows. State 4 showing the mild to strong positive FNCs in almost all networks, accounts for the smallest proportion (15%) of all windows (source data provided in Supplementary Data 2).

Table 2 showed the differences in the mean dwell time and fraction time in all 4 states between the two groups. Compared with the HC group, the MDD group had an increased fraction time (*p* = 0.003, FDR-corrected) in State 1, as well as decreased mean dwell time (*p* = 0.010, FDR-corrected) and fraction time (*p* = 0.003, FDR-corrected) in state 3. No difference in temporal properties was found in states 2 and 4 (source data provided in Supplementary Data 3).

**Table 2.** Temporal properties differences between groups.

|  | MDD group | HC group | t value | *p*-value |
| --- | --- | --- | --- | --- |
| Mean dwell time |  |  |  |  |
| State1 | 25.38±16.53 | 18.28±14.28 | 2.252 | 0.054 |
| State2 | 12.59±7.52 | 14.01±9.89 | -0.793 | 0.573 |
| State3 | 7.97±6.52 | 15.30±14.69 | -3.161 | **0.010\*** |
| State4 | 11.13±16.49 | 9.98±10.44 | 0.409 | 0.684 |
| Fraction time (%) |  |  |  |  |
| State1 | 0.51±0.22 | 0.36±0.23 | 3.302 | **0.003\*\*** |
| State2 | 0.23±0.19 | 0.25±0.23 | -0.324 | 0.747 |
| State3 | 0.12±0.12 | 0.24±0.23 | -3.504 | **0.003\*\*** |
| State4 | 0.14±0.20 | 0.15±0.19 | -0.148 | 0.883 |

All quantitative data are expressed as mean ± standard deviation; Abbreviations: MDD = major depressive disorder; HC = healthy controls. *p* values displaying significant differences between HC and MDD are marked with an asterisk, \**p* < 0.05 \*\**p* < 0.01, the significance was corrected by FDR multiple comparison correction.

### 2.4. Relationship between temporally changed states and MDD symptom severity

MDD subjects showed significantly shorter time of stay in state 3 and longer in state 1, compared to HC subjects. The correlation analysis was performed between the symptom severity measures and the temporal properties of the 4 states. In Table 3 and 4, only the mean dwell time of state 3 were significantly negatively correlated to the total HAMD scores, as well as the HAMA mental anxiety measure. However, the temporal properties of states 1, 2, 4 were not significantly correlated to HAMD and HAMA scores measures of MDD after FDR-correction (See Supplementary Table S1, S2 in Supplementary materials). All significant correlations were presented as scatter plots with trend lines in Figure 4 (source data provided in Supplementary Data 4).

**Figure 3:**
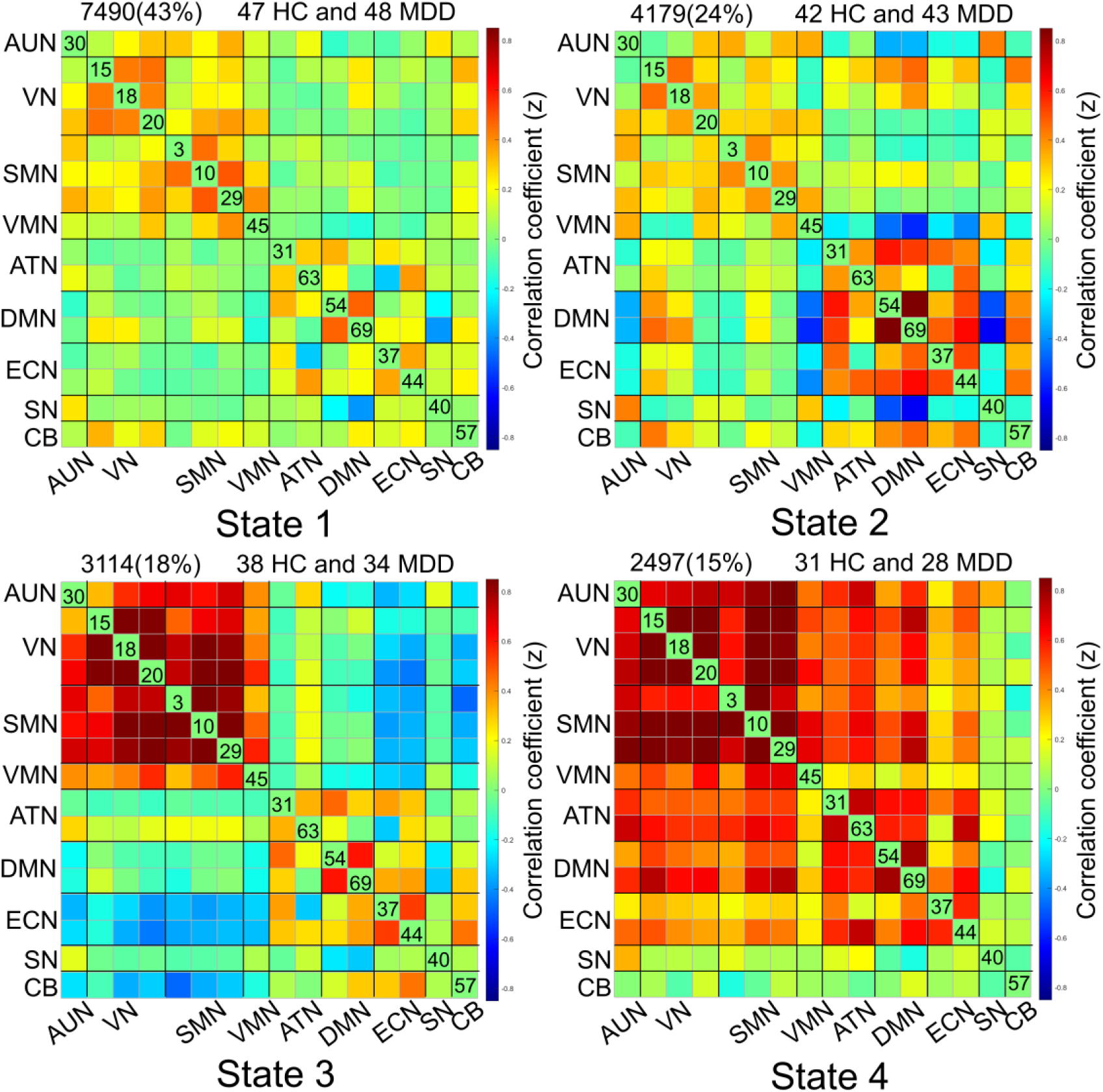
The centroids of each state. The four states were calculated and clustered by k-means method, numbers of windowed FNC and corresponding percentage have shown above the matrix, as well as the number of health control and patients that entered the state.

**Figure 4:**
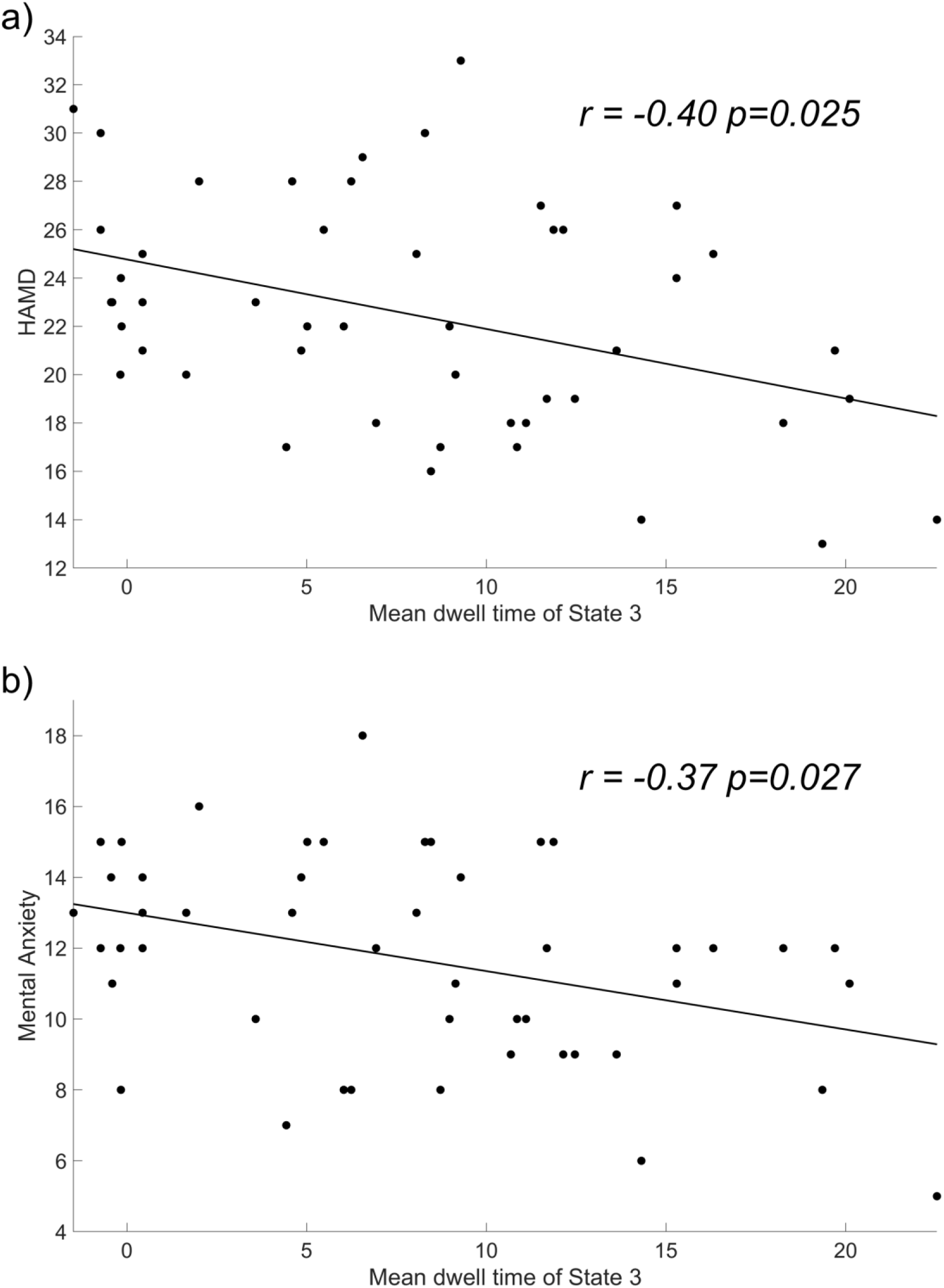
The correlation between the mean dwell time of the antagonistic state and the severity of symptoms. The correlation analysis showed that the mean dwell time of the antagonistic state in MDD was negatively related to (a) HAMD score (r = ×0.40, FDR-corrected *p* value = 0.025) and (b) the severity of the mental anxiety (r = ×0.37, FDR-corrected *p* value = 0.027).

**Table 3.** The correlation between the temporal properties of the antagonistic state and the HAMD score.

| Variables | HAMD-total score | HAMD-retardation | HAMD-cognitive<br>disorder | HAMD-sleep<br>disorders | HAMD-anxiety/so<br>matization |
| --- | --- | --- | --- | --- | --- |
| The mean dwell time of antagonistic state |  |  |  |  |  |
| R value | -0.40 | -0.06 | -0.24 | -0.23 | -0.28 |
| *p* value | **0.025\*** | 0.680 | 0.168 | 0.154 | 0.133 |
| Fraction time of antagonistic state |  |  |  |  |  |
| R value | -0.32 | 0.07 | -0.26 | -0.19 | -0.24 |
| *p* value | 0.125 | 0.638 | 0.188 | 0.245 | 0.177 |

\* $p < 0.05$ , the significance was corrected by FDR multiple comparison correction.

**Table 4.** The correlation between the temporal properties of the antagonistic state and the HAMA score.

| Variables | HAMA-total score | HAMA-Mental anxiety | HAMA-Somatic anxiety |
| --- | --- | --- | --- |
| The mean dwell time of antagonistic state |  |  |  |
| R value | -0.28 | -0.37 | -0.12 |
| *p* value | 0.075 | **0.027\*** | 0.424 |
| Fraction time of antagonistic state |  |  |  |
| R value | -0.35 | -0.40 | -0.20 |
| *p* value | 0.087 | 0.108 | 0.265 |

\* $p < 0.05$ , the significance was corrected by FDR multiple comparison correction.

## 3. Discussion

Combining sFNC and dFNC analysis, this study thoroughly investigated the FNC abnormality in MDD patients compared with HCs. We found mainly: (1) MDD patients exhibited decreased sFNC between VN and SMN and increased sFNC between VN and CB; (2) four different states were identified in dFNC analysis with MDD patients showed shorter time dwelling in state 3 (antagonistic state between executive-motor control and sensorimotor domain), and longer time dwelling in State 1 (whole brain weakly-connected state); (3) only the mean dwell time of state 3, not state 1, 2 and 4, were negatively correlated to the HAMA anxiety and HAMD scores. To the best of our knowledge, it is the first to reveal the distinct attenuated antagonism between executive-motor control function networks and sensorimotor domain in MDD, as well as its important relevance to MDD symptoms, especially in a first-episode drug-naive MDD cohort, from a dynamic functional connectivity perspective.

In sFNC analysis, the significant hypo-connectivity was detected between SMN and VN with slightly reduced within-SMN and within-VN connectivity (Figure 2c, 2d). Two mega-analysis studies have reported a hypo-connectivity within SMN [14,36], reduced regional homogeneity [37] and fractional amplitude low-frequency fluctuation (fALFF) [38] within SMN in depressed patients. In addition, decreased SMN connectivity with VN and ATN has been shown to correlate with an incapacity to distract attention from negative stimuli [39]. These results, together with our findings, may be the intuitive evidence supporting the connection between psychomotor retardation and the functional network corruption of the sensorimotor domain in brain in MDD [40]. In another word, sensorimotor cortical region involved in sensory information processing may undergo a functional decline in the pathogenesis of MDD.

The sFNC reflects the average FC over a period of time, while dFNC provides global information about FC changes over time. In a most recent study, dFNC analysis on large sample-size multi-center MDD data showed activated DMN, de-activated subcortical-cerebellum network, activated ATN, and de-activated DMN-ATN in MDD [41]. As we expected, in addition to sFNC, dFNC provided complementary information to characterize brain network dynamics [42]. In agreement with previous studies, we found in drug-naive first episode MDD patients tend to stay longer in the weakly-connected state [7,43]. This result shed light on previous sFNC findings of reduced global connectivity and lacked integration between resting-state subnetworks in MDD [11,13,44].

More importantly, this study demonstrated a new finding that the first episode MDD brains stays longer in the globally weakly-connected state (state 1), at the cost of staying less in the antagonistic state (state 3). This antagonistic state possesses distinct characteristics including: 1) the strong association within the sensorimotor domain (SMN-AUN-VN-VMN); 2) the association within cognitive-control domain (ATN-DMN-ECN) and 3) the antagonism between executive-motor control domain (ECN-CB) and sensorimotor domain. CB carries the work of motor control while ECN is the center of working memory and is also responsible for decision-making [45–47]. They are often referred as the key networks been affected in MDD but the directions of changes were sometimes not conclusive [10,41,47,48]. In the sFNC analysis in this study, the VN-CB connectivity was found increased in drug-naive first episode MDD comparing to HC. However, revealed by dFNC, this phenomenon was exactly resulted from the reduced stay in the antagonistic state and prolonged stay in globally weakly-connected state in MDD, where the VN-CB antagonistic connection was affected particularly. Our dFNC results thus provided a more in-depth understanding of the underlying alterations of FNC dynamics.

Persistent antagonism has been considered as an intrinsic property of the brain [49]. Previous rs-fMRI reports have shown spontaneous anti-correlation activity between DMN and ATN/SN, indicating that anti-correlation between brain networks is an important component of widely distributed neuroanatomical networks in the healthy population [50,51]. Also in healthy populations, stronger functional antagonism in these brain networks may facilitate processes such as the shift of attention from internal executive attention networks to the external environment, reducing the likelihood of rumination [52]. Martino et al. identified an imbalance in the interactions between DMN and SMN in depressed patients in terms of neuronal activity [53]. They hypothesized that this may lead to an excessive focus on internal thought contents at the expense of external environmental contents with inhibition in psychomotor behaviors.

Our key findings supported this hypothesis and provided more details for the theory: Whole-brain weak connected states could be associated with self-referential thought processing [54]. Normal integration of whole-brain networks is disturbed in MDD and it could be due to greater investment in self-focused rumination in the resting-state depressed patients [7,55]. The ECN and CB networks formed the executive-motor control domain and are constantly prepared to handle the information delivered from the sensorimotor domain. In the antagonistic state (state 3) found in this study, the normal brains were prepared for the processing of external sensorimotor inputs and the follow-up task-oriented control, ready to transfer the outside world information to the high-order cognitive-control domain. However, in MDD, this state was significantly weakened and reduced. This reduced antagonistic state could in turn prevent the conversion of effective cognitive processing into action and emotional expression. Therefore, the functional segregation between the cognitive-control and sensorimotor domains might also be the root cause for MDD patients tending to be more strongly involved in their inner world than healthy individuals [22,56].

Besides, we found that among all four states, the MDD symptom severity measures were only negatively correlated to the dwelling time of the antagonistic state. After evaluating the effect of illness duration on dFNC properties, we believe that this disrupted dynamics of antagonistic state is not affected by duration of illness (See Supplementary Table S3 in supplementary materials) but is rooted at the MDD onset. Given the strong evidence from both the group differences of the temporal properties of dFNCs, as well as their correlations to MDD symptoms, we cautiously propose that the reduced dwell time of the antagonistic state is the underlying core change of brain connectivity in early un-treated depression.

To be noted, several fMRI studies have also focused on FNC state dependence in MDD patients while changes in antagonistic state have not been identified. There are three possible reasons for the discrepant finding. Compared to previous studies [7,43], our subjects were restricted to the early stage of first-episode without medication, which excludes the potential effects of long disease duration and medication on brain function [31,32]. The only study of first-episode, drug-naive MDD were conducted in adolescent patients [30] while we focused on adult patients between 18–50 years old in order to avoid the potential effects of age-related neurodevelopment [57,58]. Most importantly, we also evaluated the global signal before we proceed to the dynamic fMRI analysis, which helped us to better understand the statistical underpinning of antagonistic state changes. The global signal we extracted from the fMRI time series did not show a group difference between MDD and HC (See Supplementary Figure S1 in supplementary materials, source data for Fig. S1 provided in Supplementary Data 5). Thus, we are confident that this diminished antagonistic state we find in our dynamic fMRI analysis should not be an outcome of the potential global signal shift in the early phase of depression. Thus we believe the antagonistic state could serve as an important pathophysiological marker in the early phase of MDD which changed at the onset of depression.

Currently, our cross-sectional study mainly focused on the changes in the dFNC state in the early stages of MDD without medication. Future longitudinal studies need to investigate the effects of antidepressants on important dFNC patterns, especially this antagonistic state. We note that the sample size of this study was intermediate (48 subject per group) as our recruitment was confined to first-episode, drug-naive patients with MDD. However, our sample size still enables us to find consistently significant changes in brain connectivity and is sufficient to ensure certain stability in the dFNC analysis. Overall, future studies to further investigate the underlying mechanism of the antagonism between executive-motor control networks and sensorimotor networks should require larger sample size and longitudinal follow-up investigations.

## 4. Materials and Methods

### 4.1. Subjects

Forty-eight patients with major depressive disorder were enrolled in the present study. All patients were recruited from the Department of Psychiatry at a local hospital (information will be provided after the review process). Inclusion criteria of MDD groups included (1) aged from 18 to 50 years old; (2) right-handed; (3) first episode of depression and had not yet received drug treatment;(4) the symptoms complied with the diagnostic criteria for major depression according to the Diagnostic and Statistical Manual of Mental Disorders (DSM-5) as well as the International Classification of Disease (ICD-10) diagnostic criteria. The Hamilton Depression Rating Scale (HAMD) was acquired for each patient.

A total of 48 sex- and age-matched healthy controls were recruited as a control group, and all the controls were free from neurological and psychiatric disorders. All subjects were recruited by our collaborating hospital staff. The inclusion criteria were as follows: (1) healthy adults without a history of mental illness or a family history of mental illness; and (2) HAMD score≤7. The exclusion criteria for all participants were as follows: (1) a history of severe organic brain disease or brain trauma; (2) alcohol and tobacco addiction or drug dependence; (3) pregnant women; and (4) contraindications for MRI scans.

This study was approved by the ethical committee of a local hospital according to the standards of the Declaration of Helsinki, and written informed consent was obtained from each participant (ethical approval document can be provided for review).

### 4.2. MRI data acquisition and processing

All MRI images were obtained using a GE discovery 3.0T scanner with a 16-channel-phased array head coil. The resting state fMRI images were collected using an echo-planar imaging sequence with the following parameters: TR/TE = 2000/30 ms; flip angle (FA) = 90°; field of view (FOV) = 220 ×220mm²; matrix = 64 ×64, slice thickness = 3.2mm, no gap, number of slices = 43 interleaved axial oblique; and 200 time points (6:40 min). Sagittal three-dimensional T1-weighted images (3D-T1WI) were obtained by a three-dimensional fast spoiled gradient-echo sequence with the following parameters: TR/TE=8.22ms/3.192ms, thickness=1mm, no gap, FOV=256×256mm², matrix=256×256, slices= 176. All participants have been provided with written informed consent. During the scan, each participant was asked to keep still with their eyes closed, but not to fall asleep and not to think about anything.

Functional images were preprocessed using SPM12. The main steps were as follows: (1) the first ten seconds volume were excluded to allow data to keep equilibrium; (2) slice-timing correction and motion correction were performed. Subjects revealed head motion of >1.5mm translation or >1.5°angular rotation in any direction were excluded (No participants were excluded due to head motion); (3) the scans were spatially normalized to the Montreal Neurological Institute (MNI) space, and the voxel was resized to 3mm×3mm×3mm, and the Gaussian kernel with a full-width at half maximum (FWHM) of 6 mm was used for spatial smoothing.

#### Group independent component analysis (ICA)

The ICA analyses were then performed on the preprocessed fMRI images by Group ICA of the fMRI Toolbox (GIFT, version 4.0c) (http://mialab.mrn.org/software/gift). With infomax algorithm, we chose 70 components to be extracted at first [59] and this algorithm was repeated 20 times in the ICASSO to confirm the stability of the separation of different components [60].

Each participant’s spatial maps and corresponding time-courses were provided by the GICA back reconstruction algorithm. The time-courses of each resting-state networks (RSNs) were detrended and despiked [61] and then the time-courses were filtered using a low-pass filter of 0.01∼0.15Hz. The correlation between each voxel’s time-course and the mean time-course within each IC was presented as Z-score.

Finally, 16 ICs were selected as the major networks based on the criteria listed in a previous work [62] : 1) the IC’s peak coordinates located primarily in gray matter; 2) there was no spatial overlap with vascular, ventricular and susceptibility artifacts; 3) the time-courses mainly consisted of low frequency signals. 4) the time-courses possess high dynamic range between the minimum and maximum power frequencies.

#### Static FNC (sFNC) and dynamic FNC (dFNC) analysis

The MDD and HC groups were separately calculated and averaged. The 16 ICs’ time course obtained from ICA was used to calculate sFNC correlation coefficient matrix (cc-matrix). We obtained this cc-matrix with 120 (16×15/2) edges reflecting the sFNCs between the 16 ICs and the correlation between each pair of ICs was presented as Z-score.

The sliding window method was used to evaluate dFNC. According to previous studies [63], the window was set to 15 time points (15TRs, 30s), and a Gaussian window (σ=3TRs) was set to complete the convolution, with each step advancing 1 TR, 180 windows were obtained in the end. In addition, the L1 norm was imposed in the LASSO framework with 10 repetitions to ensure the sparsity of matrix. Then we applied the K-means algorithm with the sqEuclidean distance to divide the dFNC windows into a set of separate clusters. In total 500 iterates and 150 replicates were set to avoid the local minima. Finally, we computed the dFNC properties such as mean dwell time and fraction time according to the established method [64].

### 4.3. Statistics and Reproducibility

Statistical analysis was performed using SPSS software (version 25.0). The normal distribution of the quantitative data was assessed by the Kolmogorov-Smirnov test. Between-group differences in sex ratio were determined by chi-square test, and between-group differences in other clinical variables were analyzed by two-sample t-test or Mann-Whitney U test. Additionally, we examined the group differences in sFNC and dFNC properties in each state with age, sex, and educational level as nuisance covariates. Significance was set at *p* <0.05 after false discovery rate (FDR) corrections. The correlations between temporal properties of dFNC and the MDD symptom severity such as HAMA and HAMD scores were examined by using a partial correlation analysis, adjusting for age, sex, and educational level. Further multiple comparisons correction was performed among the parallel compared measures by the Benjamini-Hochberg false discovery rate, and *p* value < 0.05 was considered statistically significant. No technical replicate was performed (i.e. each data plot comes from an independent sample).

## Acknowledgements

This study was supported by the National Natural Science Foundation of China (Grant No. 32301159), the Science Technology Department of Zhejiang Province (Grant No. 2022C03029), the Basic Public Welfare Research Program of Zhejiang Province (Grant No. LGF20H090013), the Key Research and Development Program, Ministry of Science and Technology of People’s Republic of China (Grant No. 2019YFC0121003).

## Authors’ contributions

H.Y. and M.W. designed and supervised the study. Q.G., S.C., D.Z., J.D., Y.L., Y.F and T.C. conducted fMRI scanning and clinical evaluation. Z.H. and M.W. analyzed the data. M.W. and T.C. drafted the manuscript. M.W., T.C., Z.H., X.W. and L.W.C.C. reviewed and revised the manuscript. All authors contributed to and approved the final manuscript.

## Data Availability

The data that support the findings of this study are available from the corresponding authors, Min Wang and Hong Yang, upon reasonable request. The raw data are not publicly available and only available on reasonable request due to information that could compromise the privacy of the research participants.

## Code Availability

No custom code was used in this study.

## References

[1] Ferrari, A. J., Charlson, F. J., Norman, R. E., Patten, S. B., Freedman, G., Murray, C. J., … Whiteford, H. A. (2013). Burden of depressive disorders by country, sex, age, and year: findings from the global burden of disease study 2010. PLoS Med, 10(11), e1001547. doi:10.1371/journal.pmed.1001547

[2] Belzung, C., Willner, P., & Philippot, P. (2015). Depression: from psychopathology to pathophysiology. Curr Opin Neurobiol, 30, 24–30. doi:10.1016/j.conb.2014.08.013

[3] Lu, J., Xu, X., Huang, Y., Li, T., Ma, C., Xu, G., …Zhang, N. (2021). Prevalence of depressive disorders and treatment in China: a cross-sectional epidemiological study. Lancet Psychiatry, 8(11), 981–990. doi:10.1016/S2215-0366(21)00251-0

[4] Lam, R. W., McIntosh, D., Wang, J., Enns, M. W., Kolivakis, T., Michalak, E. E., … Group, C. D. W. (2016). Canadian Network for Mood and Anxiety Treatments (CANMAT) 2016 Clinical Guidelines for the Management of Adults with Major Depressive Disorder: Section 1. Disease Burden and Principles of Care. Can J Psychiatry, 61(9), 510–523. doi:10.1177/0706743716659416

[5] Li, Z., Ruan, M., Chen, J., & Fang, Y. (2021). Major Depressive Disorder: Advances in Neuroscience Research and Translational Applications. Neurosci Bull, 37(6), 863–880. doi:10.1007/s12264-021-00638-3

[6] Hamilton, J. P., Chen, M. C., & Gotlib, I. H. (2013). Neural systems approaches to understanding major depressive disorder: an intrinsic functional organization perspective. Neurobiol Dis, 52, 4–11. doi:10.1016/j.nbd.2012.01.015

[7] Yao, Z., Shi, J., Zhang, Z., Zheng, W., Hu, T., Li, Y., … Hu, B. (2019). Altered dynamic functional connectivity in weakly-connected state in major depressive disorder. Clin Neurophysiol, 130(11), 2096–2104. doi:10.1016/j.clinph.2019.08.009

[8] Pan, F., Xu, Y., Zhou, W., Chen, J., Wei, N., Lu, S., … Huang, M. (2020). Disrupted intrinsic functional connectivity of the cognitive control network underlies disease severity and executive dysfunction in first-episode, treatment-naive adolescent depression. J Affect Disord, 264, 455–463. doi:10.1016/j.jad.2019.11.076

[9] Brakowski, J., Spinelli, S., Dorig, N., Bosch, O. G., Manoliu, A., Holtforth, M. G., & Seifritz, E. (2017). Resting state brain network function in major depression - Depression symptomatology, antidepressant treatment effects, future research. J Psychiatr Res, 92, 147–159. doi:10.1016/j.jpsychires.2017.04.007

[10] Mulders, P. C., van Eijndhoven, P. F., Schene, A. H., Beckmann, C. F., & Tendolkar, I. (2015). Resting-state functional connectivity in major depressive disorder: A review. Neurosci Biobehav Rev, 56, 330–344. doi:10.1016/j.neubiorev.2015.07.014

[11] Sun, J., Du, Z., Ma, Y., Chen, L., Wang, Z., Guo, C., … Zhao, Y. (2022). Altered functional connectivity in first-episode and recurrent depression: A resting-state functional magnetic resonance imaging study. Front Neurol, 13, 922207. doi:10.3389/fneur.2022.922207

[12] Zheng, Y., Chen, X., Li, D., Liu, Y., Tan, X., Liang, Y., … Shen, D. (2019). Treatment-naive first episode depression classification based on high-order brain functional network. J Affect Disord, 256, 33–41. doi:10.1016/j.jad.2019.05.067

[13] Luo, L., Wu, H., Xu, J., Chen, F., Wu, F., Wang, C., & Wang, J. (2021). Abnormal large-scale resting-state functional networks in drug-free major depressive disorder. Brain Imaging Behav, 15(1), 96–106. doi:10.1007/s11682-019-00236-y

[14] Javaheripour, N., Li, M., Chand, T., Krug, A., Kircher, T., Dannlowski, U., … Wagner, G. (2021). Altered resting-state functional connectome in major depressive disorder: a mega-analysis from the PsyMRI consortium. Transl Psychiatry, 11(1), 511. doi:10.1038/s41398-021-01619-w

[15] Liu, J., Mo, J. W., Wang, X., An, Z., Zhang, S., Zhang, C. Y., … Cao, X. (2022). Astrocyte dysfunction drives abnormal resting-state functional connectivity in depression. Sci Adv, 8(46), eabo2098. doi:10.1126/sciadv.abo2098

[16] Raichle, M. E. (2015). The restless brain: how intrinsic activity organizes brain function. Philos Trans R Soc Lond B Biol Sci, 370(1668). doi:10.1098/rstb.2014.0172

[17] Martino, M., Magioncalda, P., Conio, B., Capobianco, L., Russo, D., Adavastro, G., … Northoff, G. (2020). Abnormal Functional Relationship of Sensorimotor Network With Neurotransmitter-Related Nuclei via Subcortical-Cortical Loops in Manic and Depressive Phases of Bipolar Disorder. Schizophr Bull, 46(1), 163–174. doi:10.1093/schbul/sbz035

[18] Yin, Y., Wang, M., Wang, Z., Xie, C., Zhang, H., Zhang, H., … Yuan, Y. (2018). Decreased cerebral blood flow in the primary motor cortex in major depressive disorder with psychomotor retardation. Prog Neuropsychopharmacol Biol Psychiatry, 81, 438–444. doi:10.1016/j.pnpbp.2017.08.013

[19] Runia, N., Yucel, D. E., Lok, A., de Jong, K., Denys, D., van Wingen, G. A., & Bergfeld, I. O. (2022). The neurobiology of treatment-resistant depression: A systematic review of neuroimaging studies. Neurosci Biobehav Rev, 132, 433–448. doi:10.1016/j.neubiorev.2021.12.008

[20] Gong, Q., & He, Y. (2015). Depression, neuroimaging and connectomics: a selective overview. Biol Psychiatry, 77(3), 223–235. doi:10.1016/j.biopsych.2014.08.009

[21] Qiu, L., Xia, M., Cheng, B., Yuan, L., Kuang, W., Bi, F., … Gong, Q. (2018). Abnormal dynamic functional connectivity of amygdalar subregions in untreated patients with first-episode major depressive disorder. J Psychiatry Neurosci, 43(4), 262–272. doi:10.1503/jpn.170112

[22] Peng, D., Liddle, E. B., Iwabuchi, S. J., Zhang, C., Wu, Z., Liu, J., … Fang, Y. (2015). Dissociated large-scale functional connectivity networks of the precuneus in medication-naive first-episode depression. Psychiatry Res, 232(3), 250–256. doi:10.1016/j.pscychresns.2015.03.003

[23] Zhang, X., Zhu, X., Wang, X., Zhu, X., Zhong, M., Yi, J., … Yao, S. (2014). First-episode medication-naive major depressive disorder is associated with altered resting brain function in the affective network. PLoS One, 9(1), e85241. doi:10.1371/journal.pone.0085241

[24] Zhou, Y., Yu, C., Zheng, H., Liu, Y., Song, M., Qin, W., … Jiang, T. (2010). Increased neural resources recruitment in the intrinsic organization in major depression. J Affect Disord, 121(3), 220–230. doi:10.1016/j.jad.2009.05.029

[25] Tang, Y., Zhang, X., Sheng, J., Zhang, X., Zhang, J., Xu, J., … Wang, J. (2018). Elevated hippocampal choline level is associated with altered functional connectivity in females with major depressive disorder: A pilot study. Psychiatry Res Neuroimaging, 278, 48–55. doi:10.1016/j.pscychresns.2018.05.002

[26] Preti, M. G., Bolton, T. A., & Van De Ville, D. (2017). The dynamic functional connectome: State-of-the-art and perspectives. Neuroimage, 160, 41–54. doi:10.1016/j.neuroimage.2016.12.061

[27] Kaiser, R. H., Whitfield-Gabrieli, S., Dillon, D. G., Goer, F., Beltzer, M., Minkel, J., … Pizzagalli, D. A. (2016). Dynamic Resting-State Functional Connectivity in Major Depression. Neuropsychopharmacology, 41(7), 1822–1830. doi:10.1038/npp.2015.352

[28] Long, Y., Cao, H., Yan, C., Chen, X., Li, L., Castellanos, F. X., … Liu, Z. (2020). Altered resting-state dynamic functional brain networks in major depressive disorder: Findings from the REST-meta-MDD consortium. Neuroimage Clin, 26, 102163. doi:10.1016/j.nicl.2020.102163

[29] Zhang, R., Tam, S. T. S., Wong, N. M. L., Wu, J., Tao, J., Chen, L., … Lee, T. M. C. (2022). Aberrant functional metastability and structural connectivity are associated with rumination in individuals with major depressive disorder. Neuroimage Clin, 33, 102916. doi:10.1016/j.nicl.2021.102916

[30] Zheng, R., Chen, Y., Jiang, Y., Zhou, B., Li, S., Wei, Y., … Cheng, J. (2022). Abnormal dynamic functional connectivity in first-episode, drug-naive adolescents with major depressive disorder. J Neurosci Res, 100(7), 1463–1475. doi:10.1002/jnr.25047

[31] Gill, H., Puramat, P., Patel, P., Gill, B., Marks, C. A., Rodrigues, N. B., … McIntyre, R. S. (2022). The Effects of Psilocybin in Adults with Major Depressive Disorder and the General Population: Findings from Neuroimaging Studies. Psychiatry Res, 313, 114577. doi:10.1016/j.psychres.2022.114577

[32] Long, Z., Du, L., Zhao, J., Wu, S., Zheng, Q., & Lei, X. (2020). Prediction on treatment improvement in depression with resting state connectivity: A coordinate-based meta-analysis. J Affect Disord, 276, 62–68. doi:10.1016/j.jad.2020.06.072

[33] Wang, Y., Wang, C., Miao, P., Liu, J., Wei, Y., Wu, L., … Cheng, J. (2020). An imbalance between functional segregation and integration in patients with pontine stroke: A dynamic functional network connectivity study. Neuroimage Clin, 28, 102507. doi:10.1016/j.nicl.2020.102507

[34] Floegel, M., & Kell, C. A. (2017). Functional hemispheric asymmetries during the planning and manual control of virtual avatar movements. PLoS One, 12(9), e0185152. doi:10.1371/journal.pone.0185152

[35] Brovelli, A., Badier, J. M., Bonini, F., Bartolomei, F., Coulon, O., & Auzias, G. (2017). Dynamic Reconfiguration of Visuomotor-Related Functional Connectivity Networks. J Neurosci, 37(4), 839–853. doi:10.1523/JNEUROSCI.1672-16.2016

[36] Yan, C. G., Chen, X., Li, L., Castellanos, F. X., Bai, T. J., Bo, Q. J., … Zang, Y. F. (2019). Reduced default mode network functional connectivity in patients with recurrent major depressive disorder. Proc Natl Acad Sci U S A, 116(18), 9078–9083. doi:10.1073/pnas.1900390116

[37] Iwabuchi, S. J., Krishnadas, R., Li, C., Auer, D. P., Radua, J., & Palaniyappan, L. (2015). Localized connectivity in depression: a meta-analysis of resting state functional imaging studies. Neurosci Biobehav Rev, 51, 77–86. doi:10.1016/j.neubiorev.2015.01.006

[38] Xia, M., Si, T., Sun, X., Ma, Q., Liu, B., Wang, L., … Group, D. I.-M. D. D. W. (2019). Reproducibility of functional brain alterations in major depressive disorder: Evidence from a multisite resting-state functional MRI study with 1,434 individuals. Neuroimage, 189, 700–714. doi:10.1016/j.neuroimage.2019.01.074

[39] Hilland, E., Landrø, N. I., Harmer, C. J., Maglanoc, L. A., & Jonassen, R. (2018). Within-Network Connectivity in the Salience Network After Attention Bias Modification Training in Residual Depression: Report From a Preregistered Clinical Trial. Front Hum Neurosci, 12, 508. doi:10.3389/fnhum.2018.00508

[40] Northoff, G., Hirjak, D., Wolf, R. C., Magioncalda, P., & Martino, M. (2021). All roads lead to the motor cortex: psychomotor mechanisms and their biochemical modulation in psychiatric disorders. Mol Psychiatry, 26(1), 92–102. doi:10.1038/s41380-020-0814-5

[41] An, Z., Tang, K., Xie, Y., Tong, C., Liu, J., Tao, Q., … Feng, Y. (2024). Aberrant resting-state co-activation network dynamics in major depressive disorder. Transl Psychiatry, 14(1), 1. doi:10.1038/s41398-023-02722-w

[42] Zheng, W., Ge, Y., Ren, S., Ran, W., Zhang, X., Tian, W., … Wang, Z. (2020). Abnormal static and dynamic functional connectivity of resting-state fMRI in multiple system atrophy. Aging (Albany NY), 12(16), 16341–16356. doi:10.18632/aging.103676

[43] Pang, Y., Zhang, H., Cui, Q., Yang, Q., Lu, F., Chen, H., … Chen, H. (2020). Combined static and dynamic functional connectivity signatures differentiating bipolar depression from major depressive disorder. Aust N Z J Psychiatry, 54(8), 832–842. doi:10.1177/0004867420924089

[44] Zhang, L., Wu, H., Xu, J., & Shang, J. (2018). Abnormal Global Functional Connectivity Patterns in Medication-Free Major Depressive Disorder. Front Neurosci, 12, 692. doi:10.3389/fnins.2018.00692

[45] Geiger, M. J., Domschke, K., Ipser, J., Hattingh, C., Baldwin, D. S., Lochner, C., & Stein, D. J. (2016). Altered executive control network resting-state connectivity in social anxiety disorder. World J Biol Psychiatry, 17(1), 47–57. doi:10.3109/15622975.2015.1083613

[46] Wang, C., Wang, Y., Lau, W. K. W., Wei, X., Feng, X., Zhang, C., … Zhang, R. (2021). Anomalous static and dynamic functional connectivity of amygdala subregions in individuals with high trait anxiety. Depress Anxiety, 38(8), 860–873. doi:10.1002/da.23195

[47] Kang, L., Wang, W., Zhang, N., Nie, Z., Gong, Q., Yao, L., … Liu, Z. (2022). Superior temporal gyrus and cerebellar loops predict nonsuicidal self-injury in major depressive disorder patients by multimodal neuroimaging. Transl Psychiatry, 12(1), 474. doi:10.1038/s41398-022-02235-y

[48] Xu, L. Y., Xu, F. C., Liu, C., Ji, Y. F., Wu, J. M., Wang, Y., … Yu, Y. Q. (2017). Relationship between cerebellar structure and emotional memory in depression. Brain Behav, 7(7), e00738. doi:10.1002/brb3.738

[49] Kucyi, A., Daitch, A., Raccah, O., Zhao, B., Zhang, C., Esterman, M., … Parvizi, J. (2020). Electrophysiological dynamics of antagonistic brain networks reflect attentional fluctuations. Nat Commun, 11(1), 325. doi:10.1038/s41467-019-14166-2

[50] Fox, M. D., Snyder, A. Z., Vincent, J. L., Corbetta, M., Van Essen, D. C., & Raichle, M. E. (2005). The human brain is intrinsically organized into dynamic, anticorrelated functional networks. Proc Natl Acad Sci U S A, 102(27), 9673–9678. doi:10.1073/pnas.0504136102

[51] Fransson, P. (2005). Spontaneous low-frequency BOLD signal fluctuations: an fMRI investigation of the resting-state default mode of brain function hypothesis. Hum Brain Mapp, 26(1), 15–29. doi:10.1002/hbm.20113

[52] Rostami, S., Borjali, A., Eskandari, H., Rostami, R., Scalabrini, A., & Northoff, G. (2022). Slow and Powerless Thought Dynamic Relates to Brooding in Unipolar and Bipolar Depression. Psychopathology, 55(5), 258–272. doi:10.1159/000523944

[53] Martino, M., Magioncalda, P., Huang, Z., Conio, B., Piaggio, N., Duncan, N. W., … Northoff, G. (2016). Contrasting variability patterns in the default mode and sensorimotor networks balance in bipolar depression and mania. Proc Natl Acad Sci U S A, 113(17), 4824–4829. doi:10.1073/pnas.1517558113

[54] Marusak, H. A., Calhoun, V. D., Brown, S., Crespo, L. M., Sala-Hamrick, K., Gotlib, I. H., & Thomason, M. E. (2017). Dynamic functional connectivity of neurocognitive networks in children. Hum Brain Mapp, 38(1), 97–108. doi:10.1002/hbm.23346

[55] Berman, M. G., Misic, B., Buschkuehl, M., Kross, E., Deldin, P. J., Peltier, S., … Jonides, J. (2014). Does resting-state connectivity reflect depressive rumination? A tale of two analyses. Neuroimage, 103, 267–279. doi:10.1016/j.neuroimage.2014.09.027

[56] Hamilton, J. P., Furman, D. J., Chang, C., Thomason, M. E., Dennis, E., & Gotlib, I. H. (2011). Default-mode and task-positive network activity in major depressive disorder: implications for adaptive and maladaptive rumination. Biol Psychiatry, 70(4), 327–333. doi:10.1016/j.biopsych.2011.02.003

[57] Wierenga, L. M., Bos, M. G. N., Schreuders, E., Vd Kamp, F., Peper, J. S., Tamnes, C. K., & Crone, E. A. (2018). Unraveling age, puberty and testosterone effects on subcortical brain development across adolescence. Psychoneuroendocrinology, 91, 105–114. doi:10.1016/j.psyneuen.2018.02.034

[58] Lockwood, K. A., Alexopoulos, G. S., & van Gorp, W. G. (2002). Executive dysfunction in geriatric depression. Am J Psychiatry, 159(7), 1119–1126. doi:10.1176/appi.ajp.159.7.1119

[59] Bell, A. J., & Sejnowski, T. J. (1995). An information-maximization approach to blind separation and blind deconvolution. Neural Comput, 7(6), 1129–1159. doi:10.1162/neco.1995.7.6.1129

[60] Himberg, J., Hyvarinen, A., & Esposito, F. (2004). Validating the independent components of neuroimaging time series via clustering and visualization. Neuroimage, 22(3), 1214–1222. doi:10.1016/j.neuroimage.2004.03.027

[61] Calhoun, V. D., Adali, T., Pearlson, G. D., & Pekar, J. J. (2001). A method for making group inferences from functional MRI data using independent component analysis. Hum Brain Mapp, 14(3), 140–151. doi:10.1002/hbm.1048

[62] Allen, E. A., Erhardt, E. B., Damaraju, E., Gruner, W., Segall, J. M., Silva, R. F., … Calhoun, V. D. (2011). A baseline for the multivariate comparison of resting-state networks. Front Syst Neurosci, 5, 2. doi:10.3389/fnsys.2011.00002

[63] Allen, E. A., Damaraju, E., Plis, S. M., Erhardt, E. B., Eichele, T., & Calhoun, V. D. (2014). Tracking whole-brain connectivity dynamics in the resting state. Cereb Cortex, 24(3), 663–676. doi:10.1093/cercor/bhs352

[64] Kim, J., Criaud, M., Cho, S. S., Diez-Cirarda, M., Mihaescu, A., Coakeley, S., … Strafella, A. P. (2017). Abnormal intrinsic brain functional network dynamics in Parkinson’s disease. Brain, 140(11), 2955–2967. doi:10.1093/brain/awx233

